# Altered food liking in depression is driven by macronutrient composition

**DOI:** 10.1101/2024.09.09.24313298

**Authors:** Lilly Thurn, Corinna Schulz, Diba Borgmann, Johannes Klaus, Sabine Ellinger, Martin Walter, Nils B. Kroemer

## Abstract

Major depressive disorder (MDD) is characterized by changes in appetite and body weight as well as blunted reward sensitivity (“anhedonia”). However, it is not well understood which mechanisms are driving changes in reward sensitivity, specifically regarding food. Here, we used a sample of 117 participants (54 patients with MDD; 63 healthy control participants, HCP) who completed a food cue reactivity (FCR) task with ratings of wanting and liking for 60 food and 20 non-food items. To evaluate which components of the food may contribute to altered ratings in depression, we tested for associations with macronutrients of the depicted items. In line with previous studies, we found reduced ratings of food wanting (*p* = .003), but not liking (*p* = .23) in patients with MDD compared to matched HCPs. Adding macronutrient composition to the models of wanting and liking substantially improved their fit (*p*s < .001). Compared to carbohydrate-rich foods, patients with MDD reported lower liking and wanting ratings for high-fat and high-protein foods. Moreover, patients with MDD showed weaker correlations in their preferences for carbohydrate-versus fat- or protein-rich foods (*p*s < .001), pointing to potential disturbances in metabolic signaling. To conclude, our results suggest that depression-related alterations in food reward ratings are more specific to the macronutrient composition of the food than previously anticipated, hinting at disturbances in gut-brain signaling. These findings raise the intriguing question whether interventions targeting the gut could help normalize aberrant reward signals for foods rich in fat or protein.

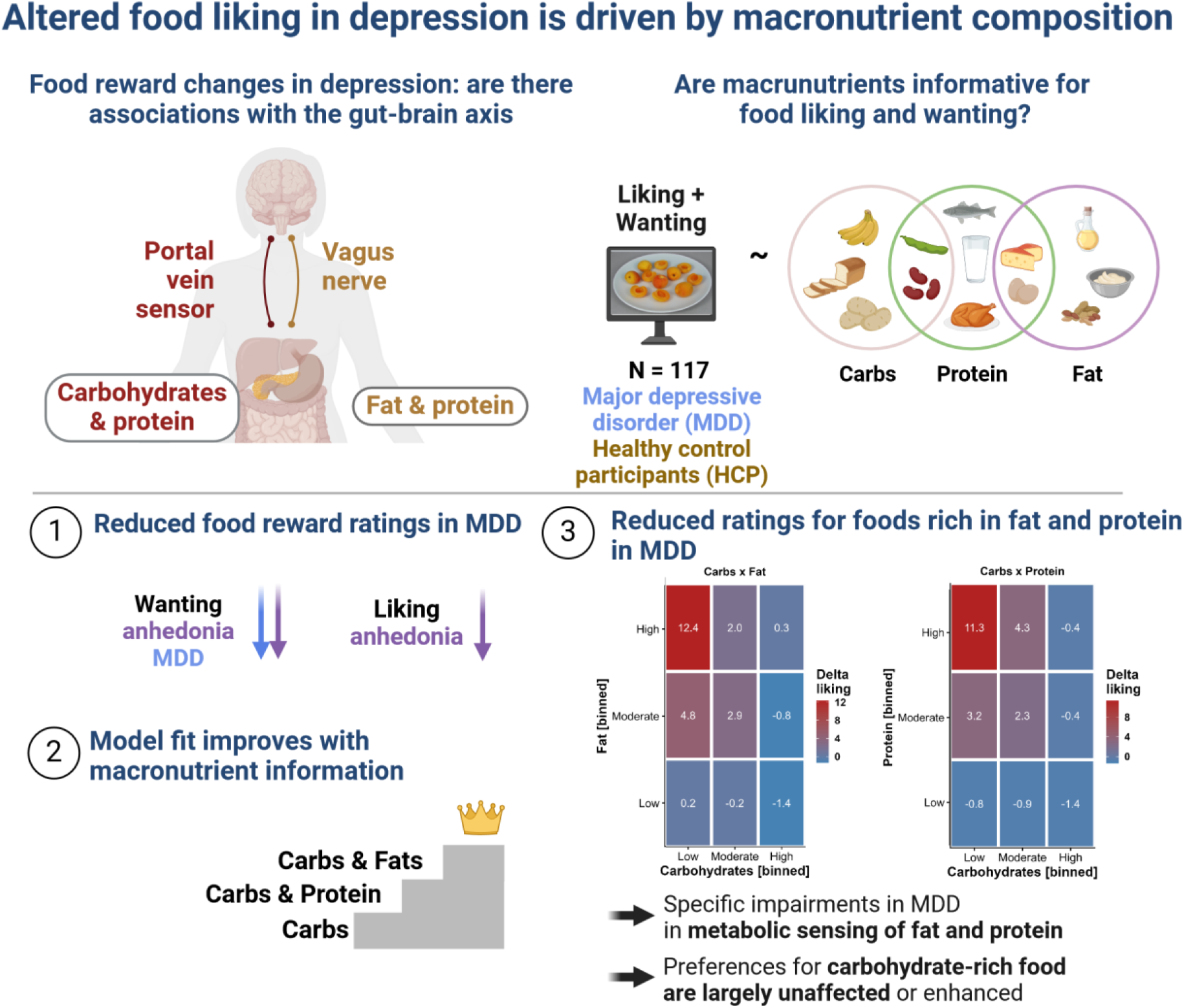

## Introduction

Major depressive disorder (MDD) affects about 280 million people worldwide (Institute of Health Metrics and Evaluation, 2023). It is characterized by diverse symptoms (Fried and Nesse, 2015a; Fried et al., 2022) such as depressed mood and loss of interest or pleasure (“anhedonia”) as cardinal symptoms. MDD also includes opposing somatic symptoms, such as increased versus decreased appetite and body weight, during a depressive episode (Polivy and Herman, 1976; Milaneschi et al., 2019). Although appetite-related symptoms are often not directly treated and may even worsen with antidepressant medication (Kazes et al., 1994; Himmerich et al., 2015), they are comparatively stable across episodes (Stunkard et al., 1990) and possibly reflect mechanistically distinct subtypes of depression (Penninx et al., 2013; Cuijpers et al., 2017; Milaneschi et al., 2017a; Milaneschi et al., 2017b; Simmons et al., 2020). Consequently, such opposing changes in appetite and body weight have been associated with differential brain responses to food cues (Simmons et al., 2016) and distinct functional connectivity profiles of the reward circuit (Kroemer et al., 2022). These emerging results suggest that an improved phenotyping of appetite-related symptoms in depression promises a more refined diagnosis of reward dysfunction (Fried et al., 2014; Fried and Nesse, 2015b; Schulz et al., 2024). In the long run, such insights may improve the allocation of suitable treatments since effective therapies for somatic symptoms of MDD should conceivably consider the direction of changes in appetite and body weight.

To better characterize appetite-related processes and symptoms, many studies use food cue reactivity (FCR) paradigms (van der Laan et al., 2011; Smeets et al., 2012). During these FCR tasks, participants view various food images while concurrent brain (Hare et al., 2009; Frank et al., 2010; Guthoff et al., 2010; Kroemer et al., 2013a; Kroemer et al., 2013b; Charbonnier et al., 2018) or physiological responses (Rebollo et al., 2021) are collected, or ratings scales are embedded to capture cue-elicited responses guiding eating behavior (e.g., taste, healthiness, wanting, or liking; (Charbonnier et al., 2015; Müller et al., 2022). Food images are typically contrasted against non-food images (Koepp et al., 2021; Müller et al., 2022), or grouped into categories, for example, based on high versus low energy density (Dimitropoulos et al., 2012; Mehta et al., 2012). Recent studies have demonstrated that FCR tasks help reveal nuanced information about the encoding of food reward by considering the macronutrient composition of the depicted food (Small and DiFeliceantonio, 2019; Motoki et al., 2021). For example, DiFeliceantonio et al. (2018) have shown supra-additive effects of fat and carbohydrate content on food reward using brain responses and willingness to pay as outcomes. These findings extrapolate previous work on the correspondence between willingness to pay and the true energy density of food, but not the perceived energy content (Tang et al., 2014). Such nutritional information is estimated surprisingly quickly as the accuracy of ratings does not improve if participants have more time than 100 ms to evaluate the food (Motoki et al., 2021). Therefore, conventional FCR tasks may provide useful insights about altered food reward processing of macronutrients in depression, even though a comprehensive assessment is lacking to date.

Associating the macronutrient composition of food with markers of reward dysfunction in depression is also promising in light of plausible neurobiological links with disturbed energy metabolism (Kenny, 2011; Milaneschi et al., 2019; Borgmann and Fenselau, 2024). Relating subjective reinforcing values of food to nutritive values of its fat and carbohydrates content relies on parallel neural afferent signaling pathways (Small and DiFeliceantonio, 2019; de Araujo et al., 2020; McDougle et al., 2024). Whereas fat-induced reward signals are mainly transmitted via vagal afferents (Tellez et al., 2013; de Araujo, 2016; de Araujo et al., 2020; Goldstein et al., 2021), carbohydrate-induced reward signals are primarily conveyed via portal vein sensors and spinal afferents (Goldstein et al., 2021; Bacharach et al., 2023) although vagal afferents contribute as well (Fernandes et al., 2020). For protein-induced effects, there is mostly indirect evidence pointing to vagal afferent signals (de Lartigue and Diepenbroek, 2016), although subdiaphragmatic vagotomies in rats show that it is not an obligatory pathway for the transfer of energy-related information to the brain (L’Heureux-Bouron et al., 2003). Moreover, distinct gut-innervating sensory neurons differentially control feeding and glucoregulatory neurocircuits, potentially pointing to specific targets for improved metabolic control (Bai et al., 2019; Tan et al., 2020; Borgmann et al., 2021; Li et al., 2022).

Enhanced craving for carbohydrates has often been observed in patients with MDD with an alleged link to increased tryptophan metabolism and brain serotonin levels (Fernstrom and Wurtman, 1971; Fernstrom et al., 1987; Wurtman and Wurtman, 1995; Markus, 2007). Accordingly, self-reported differences in macronutrient consumption are associated with a risk for depression in large cross-sectional studies (Cheng et al., 2024) and Mendelian randomization studies even suggest a protective effect of carbohydrate intake (Yao et al., 2022). Since depression is often comorbid with obesity (Milaneschi et al., 2019) and type 2 diabetes (Lustman et al., 2000; Anderson et al., 2001; Geraets et al., 2020), altered energy metabolism could contribute to reward deficits and somatic symptoms (Pistis et al., 2021) and an improved knowledge of the pathways contributing to specific depressive symptoms may help identify suitable therapies. Illustratively, recent studies have demonstrated that patients with MDD show altered lipid metabolism (Tkachev et al., 2023) and targeting vagal afferent projections with vagus nerve stimulation could potentially alleviate key symptoms of depression (Edwin Thanarajah and Reif, 2022; Ferstl et al., 2024; Teckentrup and Kroemer, 2024), including food reward deficits (Bodenlos et al., 2007; Han et al., 2018; Koepp et al., 2021; McDougle et al., 2024). In sum, evaluating whether altered appetite in depression is specifically correlated with the macronutrient composition of food may reveal physilogically relevant insights.

To address the pressing question whether altered food reward in depression is associated with the macronutrient composition of the depicted food, we investigated patients with MDD and healthy control participants (HCPs) using a conventional FCR task with 60 food and 20 non-food images (Koepp et al., 2021; Müller et al., 2022). In addition, we characterized changes in appetite and body weight in patients with MDD using the SIGH-ADS interview (Williams and Terman, 2003) that covers atypical symptoms of depression. Crucially, we also collected fasting levels of glucose, insulin, and triglycerides to estimate individual differences in insulin sensitivity as measures of metabolic disturbances. To evaluate associations with the macronutrient composition of food, we used linear mixed-effects models predicting wanting and liking ratings using the true nutritional value of the rated food images (Charbonnier et al., 2015). By demonstrating specific associations of reward deficits in depression with foods rich in fat, but not carbohydrates, our study hints at the potential of incorporating macronutrient information into the research of somatic symptoms.

## Methods

### Participants

As part of an ongoing cross-sectional study, we analyzed a sample of 117 participants (54 diagnosed with MDD and 63 healthy control participants, HCP). The reported results are part of a larger preregistered study about the effects of the gut-brain axis on depression (https://clinicaltrials.gov/study/NCT05318924, Fahed et al. (2024), Schulz et al. (2024)). On average, participants had a mean age of 30 ± 8 years and a mean body mass index (BMI) of 23.6 ± 3.2 kg/m^2^ (see Table 1). Compared to the group of patients with MDD, we oversampled older HCPs for an additional part of the study involving PET/fMRI. To avoid confounding effects of age, we accounted for age in all mixed-effects analyses (with additional confounds).

**Table 1:** Sample characteristics.

| Characteristic | Overall<br>N = 117<sup>1</sup> | HCP<br>N = 63<sup>1</sup> | MDD<br>N = 54<sup>1</sup> | p-value<sup>2</sup> |
| --- | --- | --- | --- | --- |
| Age, years | 30.1 ± 7.7 | 31.9 ± 7.6 | 28.1 ± 7.3 | .007 |
| Females | 71 (61%) | 38 (60%) | 33 (61%) | >.9 |
| Weight, kg | 71.6 ± 13.2 | 72.3 ± 12.4 | 70.8 ± 14.1 | .5 |
| Height, m | 1.74 ± 0.09 | 1.74 ± 0.08 | 1.73 ± 0.11 | .5 |
| Hip circumference, cm | 96.3 ± 11.4 | 96.4 ± 12.4 | 96.1 ± 10.2 | .9 |
| Missing | 1 | 0 | 1 |  |
| Waist circumference, cm | 81.4 ± 11.4 | 80.2 ± 11.1 | 82.8 ± 11.8 | .2 |
| Missing | 1 | 0 | 1 |  |
| Waist-to-Hip Ratio | 0.85 ± 0.11 | 0.83 ± 0.08 | 0.87 ± 0.14 | .11 |
| Missing | 1 | 0 | 1 |  |
| BMI, kg/m<sup>2</sup> | 23.6 ± 3.2 | 23.7 ± 3.0 | 23.5 ± 3.5 | .7* |

<sup>1</sup>Mean ± SD; n (%)<sup>2</sup>Two Sample *t*-test or Pearson's Chi-squared test

All participants were screened for eligibility prior to investigation by phone. Individuals were excluded when over the age of 50 or younger than 20 or if their BMI exceeded 30.0 kg/m^2^ or was below 18.5 kg/ m^2^. Furthermore, they were excluded if they had the comorbid disorder schizophrenia, bipolar disorder, severe substance abuse, or a severe neurological condition. HCPs were excluded if a mood or anxiety disorder was present. Due to high comorbidities with anxiety disorders (Kaufman and Charney, 2000), this criterion was not applied to patients with MDD. Individuals were also excluded if they suffered from one of the following conditions within 12 months before screening: eating disorder, obsessive-compulsive disorder, unprocessed trauma and stressor-related disorder, and somatic symptom disorder. Moreover, they were screened for medication or illnesses influencing body weight and current pregnancy or nursing were additional exclusion criteria for women. Assignment to the MDD group required having MDD at the time of screening (i.e., within 3 months of enrollment) according to DSM-5 criteria. To generalize results to typical patients with MDD, there was no restriction concerning the type of therapy or medication except for changes of medication type or dosage within the preceding two months.

Recruitment took place at the Department of Psychiatry and Psychotherapy, University of Tübingen, Germany, and on social media using online advertisements and flyers. Participants were asked to provide written consent prior to the first session. Compensation was given in the form of monetary and food-related rewards (snacks) resulting from the performance in decision-making games. Furthermore, participants received €50 or 5 credit points (if they were students and selected them) for full study completion. The study was approved by the institutional review board that is the local ethics committee of the University of Tübingen, Faculty of Medicine, in accordance with the Declaration of Helsinki (as revised in 2008).

### Experimental procedure

The study was conducted at the Department of Psychiatry and Psychotherapy in Tübingen, Germany, and consisted of two parts, completed either on one or two separate days. Participants gave written informed consent before beginning the study. Participation included an online assessment with questionnaires, for example, to assess symptoms of depression [Becks Depression Inventory, BDI-II (Hautzinger et al., 2009)] and anhedonia [Snaith-Hamilton Please Scale in German, SHAPS-D (Franz et al., 1998)]. During the first session, we conducted the Structured Clinical Interview for DSM-5 (SCID) (First, 2014). In addition, we used the Structured Interview Guide for the Hamilton Rating Depression Scale with Atypical Depression Supplement (SIGH-ADS) (Williams & Terman, 2003) within the MDD group to calculate delta appetite scores (>0 increase; <0 decrease) that reflect opposing appetite-related symptoms (Kroemer et al., 2022; Schulz et al., 2024). As this part took approximately 30 min for HCP and up to 2 h for patients with MDD, HCPs were offered to complete both parts within one day, whereas patients with MDD were invited to come for the second part on another day. The second part consisted of blood draws and a battery of reward-related tasks. On this day, participants were investigated after an overnight fast (∼12 h) in which they were only allowed to consume unsweetened beverages (e.g., water, tea). First, participants had to answer questions in their current subjective metabolic and affective state using visual analogue scales (VAS; Fahed et al. (2024)). Second, fasting blood samples were taken to determine hormone levels. The experimenters also determined the participants’ height, weight, waist and hip circumference, and documented menstrual cycle (if applicable), and time of the last meal and drink. After receiving task-specific instructions, they were asked to complete a battery of reward-related tasks (Schulz et al., 2024), including an FCR task (Müller et al., 2022). The duration of the whole session was about 3.5 h and the FCR task was the first task, taking up about 20 min. Participants were provided water ad libitum. At the end of the session, they received their acquired food rewards and financial compensation.

### Food cue reactivity task

To assess anticipatory aspects of food reward, we used an FCR task (Müller et al., 2022). The task consisted of standardized images by Charbonnier et al. (2016) optimized for visual characteristics (homogenous plate with gray background). From this set of images, 60 food images and 20 non-food images depicting office supplies, serving as a control condition, were drawn (see Table S.x). The task consisted of cue and rating phases. Participants were shown one image before they were asked to evaluate them either based on (a) how much they like the item based on past experiences (liking) or (b) how much they want to receive the depicted item (wanting). Both answers were given on VAS, however, different orientations emphasized the difference in concepts. Liking was rated on a vertical scale (Lim et al., 2009) ranging from -100 (strongest disliking imaginable) to +100 (strongest liking imaginable), whereas wanting was rated on a horizontal scale ranging from 0 (not wanted at all) to 100 (strongly wanted). Each trial started with a black fixation cross on a white screen for one second, followed by an (non-)food image for 2 s. Afterwards, another fixation cross was shown prior to the rating scale, which was presented for up to 2.8 s to induce fast decision making in participants. The participants could choose their answer using an XBox controller joystick (Microsoft Corporation, Redmond, WA) and confirm by pressing A, after which a new trial started. To assess liking and wanting separately, all participants viewed the images twice. The order of stimulus presentation and rating was pseudo-randomized. The tasks were completed on a laptop (Lenovo L580) and executed using Psychtoolbox v3.0.18 (Kleiner et al., 2007) in MATLAB (2021b).

## Data analysis

### Liking and wanting ratings

To evaluate the associations of liking and wanting ratings with macronutrients, we conducted linear mixed-effects models. These models used either liking or wanting as outcome and included the centered predictors group (HCP vs. MDD), BMI, age, and sex (female vs. male). Since we used model comparisons in the next step, these base models (M0) were fitted with full maximum likelihood estimation with random intercepts and slopes. Then, we tested three extended models including macronutrient content of the depicted food (z-standardized to ensure convergence) as predictors. The first extended model (i.e., carbohydrate model, eM1) contained the term carbohydrate content. The second extended model contained the term carbohydrate content and fat content as well as their interaction (i.e., Carbohydrate × Fat model, eM2a). The third extended model contained the term carbohydrate content and protein content as well as their interaction (i.e., Carbohydrate × Protein model, eM2b). The latter two extended models were used as alternative models and compared to the base model as well as the carbohydrate model to ensure nested model comparisons. A potentially larger model including all macronutrients was not feasible with all random slopes for the interactions and would likely require more items for robust estimation of the interaction terms. We also evaluated the incremental effect of anhedonia symptoms by adding group-centered SHAPS scores as a predictor. To compare slopes within the linear-mixed effects models, we used the R function contest with a *t*-contrast vector comparing the interaction terms of group with carbohydrate, fat, and protein content. To calculate unbiased slope estimates, we ran separate models with restricted maximum likelihood estimation and only including the macronutrient content (z-standardized) as well as the interactions of Carbohydrates × Fat, or Carbohydrates × Protein, respectively. These models provide unbiased estimates because they did not include group or demographic variables as predictors while random intercepts and slopes capture individual differences from the group average (extracted with the coef function).

### Fasting blood levels

As proxies of insulin sensitivity or resistance, we calculated the homeostasis model assessment of insulin resistance (HOMA-IR) using fasting glucose and insulin levels (Matthews et al., 1985; (insulin [pmol/l] /6,945) * glucose [mg/dl] / 405), as well as the triglyceride–glucose index using fasting triglycerides and glucose levels (Unger et al., 2014; Ln(triglycerides [mg/dl] * glucose [mg/dl]) / 2). The samples were analyzed by the Central Laboratory of the Institute of Clinical Chemistry and Pathobiochemistry of the University Hospital Tübingen. The concentrations of both acylated and unacetylated ghrelin was determined in plasma using ELISA kits (#A05306 and #A05319; both from Bertin Bioreagent, Bertin Technologies, Montigny-le-Bretonneux, France; distributed by BioCat, Germany) at the Institute of Nutritional and Food Sciences, Human Nutrition, University of Bonn. To improve the distribution of the obtained blood levels, we used natural log transformation and inspected them using Q-Q plots, indicating that they were well approximated by normal distributions (Schulz et al., 2024).

### Statistical threshold and software

All statistical analysis and visualizations were performed using R version 4.3.2 (R Core Team, 2023). The linear mixed-effects models were computed using Satterthwaite’s method with the R package lmertest (Kuznetsova et al., 2017). The significance threshold was set to *p* < .05.

## Results

### Patients with MDD report lower food wanting

To evaluate group differences in food reward ratings, we first used two basal mixed-effects models as references for model comparisons. We predicted either liking or wanting using group and nuisance variables (i.e., BMI, age, sex) as predictors (grand mean centered). Patients with MDD showed no differences in food liking (*b* = -1.68, *p* = .23), but reported lower food wanting (*b* = -6.22, *p* = .003) compared to HCPs. Adding group-centered SHAPS scores as a predictor revealed that associations with food liking (*b* = -0.28, *p* = .010) and food wanting (*b* = -0.68, *p* < .001) were much more pronounced for participants that reported more symptoms of anhedonia.

### Adding the macronutrient content of food items substantially improves the model fit of liking ratings

Next, we examined whether adding the carbohydrate content (including an interaction with group with a random slope to capture inter-individual variability) of the depicted food improved the fit of the base models. For food liking, modeling the food’s carbohydrate content improved fit considerably, *χ*^2^(4) = 300.57, *p* < .001, ΔBIC = 266. Increasing the carbohydrate content by 1 SD unit (∼26.5 g/100g) led to a reduction in liking of *b* = -2.28 (*p* < .001). This association was non-significantly attenuated in patients with MDD (*b* = 0.91, *p* = .23).

We then added the food’s content of fat (including interactions with group and carbohydrate content with random slopes for all terms) and observed a considerable improvement compared to modeling only the carbohydrate content, *χ*^2^(11) = 377.51, *p* < .001, ΔBIC = 280. Increasing the fat content by 1 SD unit (∼13.4 g/100g) led to a non-significant slight reduction in liking of *b* = -0.36 (*p* = .38). This association with fat was non-significantly amplified in patients with MDD (*b* = -1.51, *p* = .071) and after accounting for the fat content, patients with MDD reported higher liking for foods with a high content of carbohydrates (*b* = 1.75, *p* = .007). A contrast comparing the slopes for carbohydrates and fat revealed a strong relative preference for carbohydrates in patients with MDD (vs. HCPs) compared to fat, *t*(117) = 3.44, *p* < .001. Fat also enhanced the liking of foods that were rich in carbohydrates, which was reflected in a strong interaction (*b* = 2.25, *p* < .001). Crucially, we also observed a significant interaction of group with the term Carbohydrates × Fat, reflecting that patients with MDD reported higher liking (*b* = 1.64, *p* = .015; Figure 1a).

**Figure 1:**
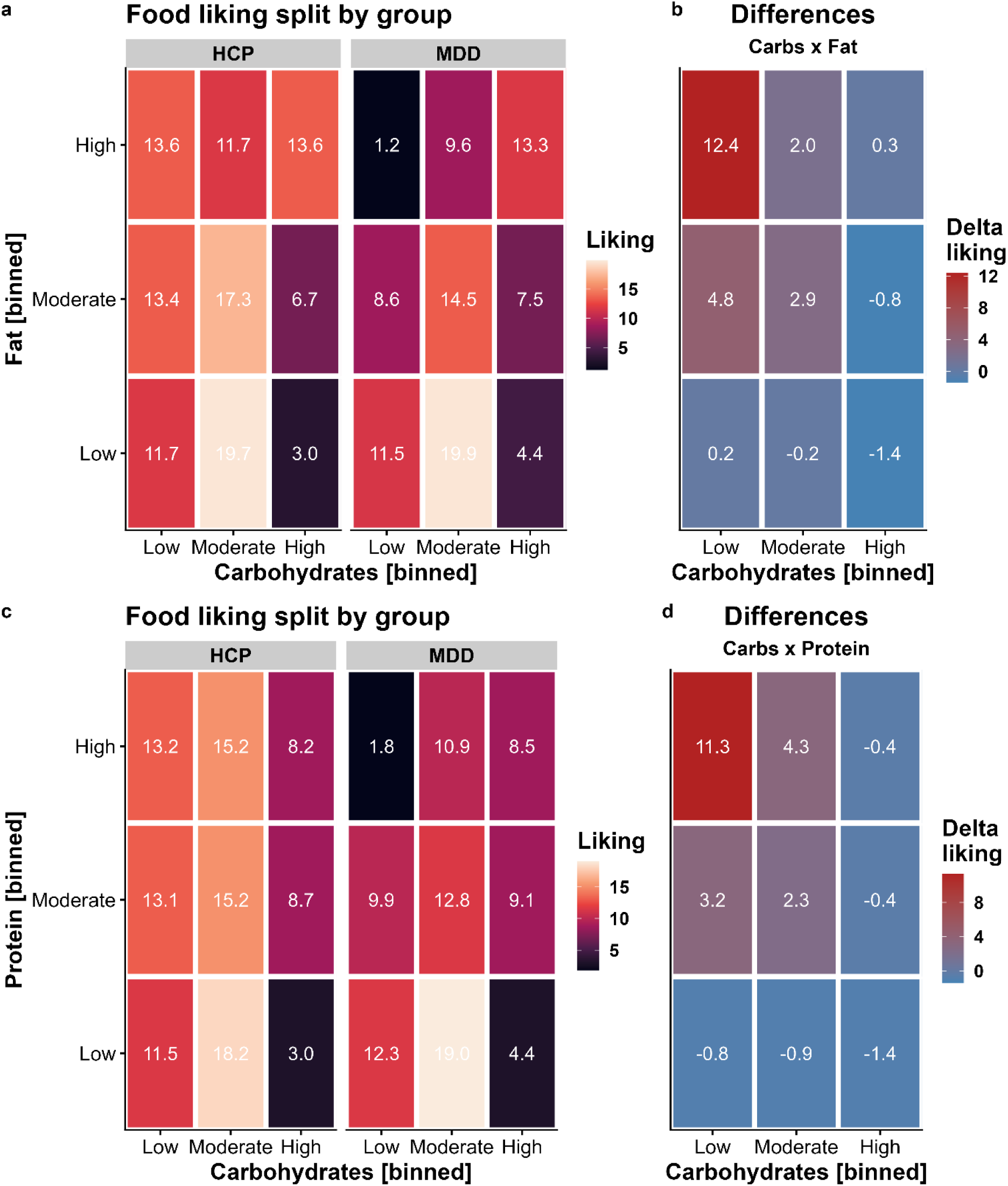
Patients with major depressive disorder (MDD) report lower liking for foods with high concentrations of fat and protein unless they contain carbohydrates as well. A: Liking ratings split by group and the macronutrients carbohydrates and fat. For display, we binned macronutrient contents so that low corresponds to the lower quartile, moderate to the second and third quartile, and high to the upper quartile. B: Differences between healthy control participants (HCP) and patients with MDD for foods according to their carbohydrate and fat content. Patients with MDD report lower liking for high-fat foods (interaction contrast: *p* < .001). C: Liking ratings split by group and the macronutrients carbohydrates and protein. D: Similar to fat, patients with MDD report lower liking for high-protein foods (interaction contrast: *p* = .003).

Alternatively, we evaluated the interaction of carbohydrates with the protein content of foods. The addition of protein instead of fat led to a smaller improvement in model fit compared to modeling the content of carbohydrates and fat, *χ*^2^(11) = 271.62, *p* < .001, ΔBIC = 174 (with the carbohydrate model as reference). Increasing the protein content by 1 SD unit (∼6.0 g/100g) led to a reduction in liking of *b* = -0.91 (*p* = .027). This association with protein was amplified in patients with MDD (*b* = -1.86, *p* = .024). A contrast comparing the slopes for carbohydrates and protein revealed a strong relative preference for carbohydrates in patients with MDD compared to protein and HCPs, *t*(117) = 3.06, *p* = .003. Similar to fat, protein also enhanced the liking of foods rich in carbohydrates, leading to a significant interaction term (*b* = 0.84, *p* = .002). However, patients with MDD did not show an enhanced interaction effect in contrast to fat (*b* = 0.55, *p* = .31; Figure 1b).

### Wanting ratings similarly reflect the macronutrient content of food

For food wanting, modeling the food’s carbohydrate content improved fit comparable to food liking, *χ*^2^(4) = 268.91, *p* < .001, ΔBIC = 233. Increasing the carbohydrate content by 1 SD unit (∼26.5 g/100g) led to a reduction in wanting of *b* = -2.72 (*p* < .001). This association was non-significantly attenuated in patients with MDD (*b* = 0.75, *p* = .52).

Analogous to liking, adding the food’s fat content led to a considerable improvement compared to modeling only the carbohydrate content for wanting, *χ*^2^(11) = 422.20, *p* < .001, ΔBIC = 325. Increasing the fat content by 1 SD unit (∼13.4 g/100g) led to a non-significant reduction in wanting of *b* = -1.17 (*p* = .058). This association with fat was non-significantly amplified in patients with MDD (*b* = -1.54, *p* = .21). A contrast comparing the slopes for carbohydrates and fat revealed a moderate relative preference for carbohydrates compared to fat in patients with MDD versus HCPs, *t*(117) = 2.38, *p* = .019. Fat also enhanced the wanting of foods rich in carbohydrates, which was reflected in a strong interaction (*b* = 3.75, *p* < .001). We also observed a non-significant interaction of group with the term Carbohydrates × Fat, reflecting that patients with MDD reported marginally higher wanting (*b* = 1.96, *p* = .052; Figure 2a) that was numerically comparable to liking.

**Figure 2:**
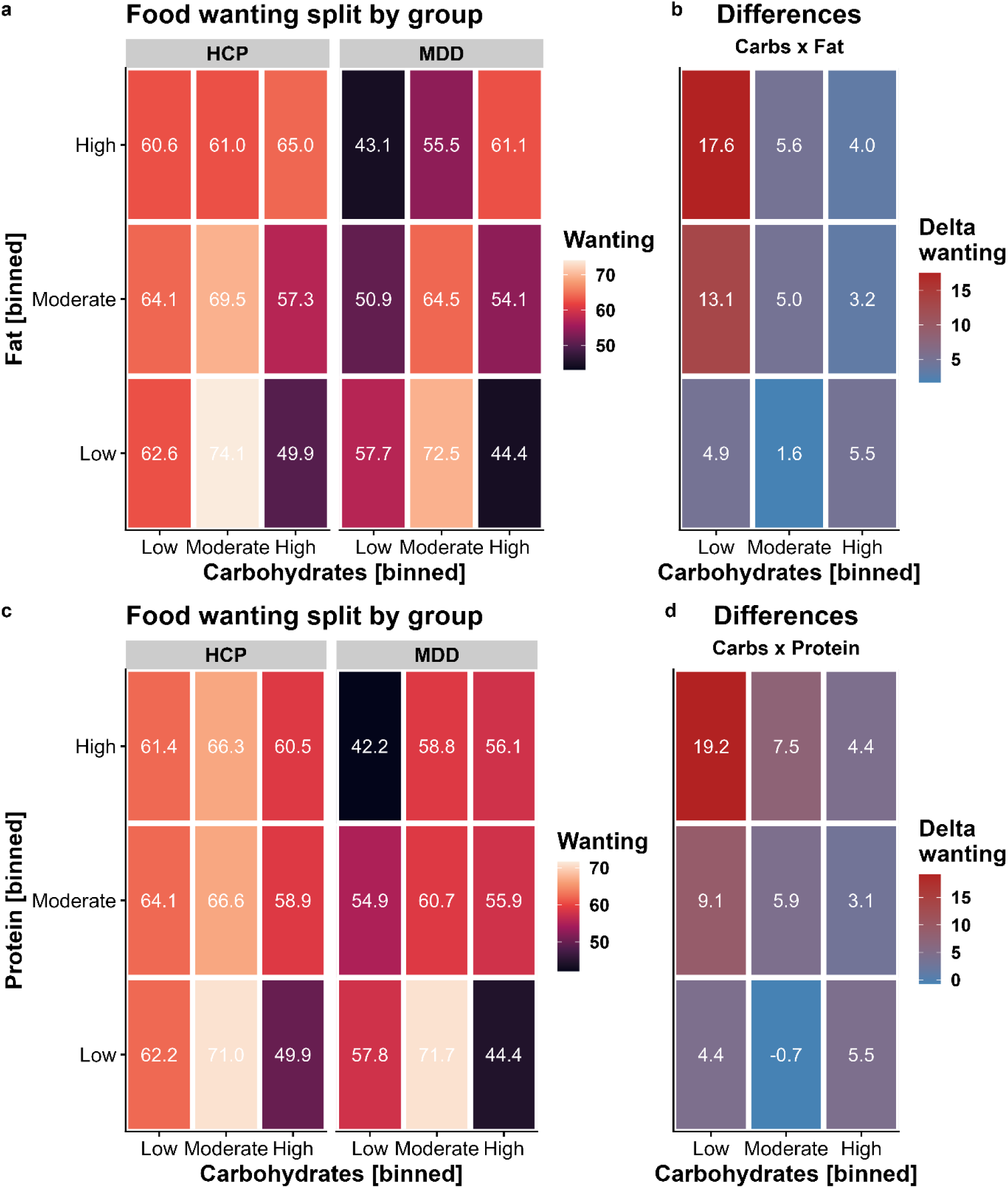
Patients with major depressive disorder (MDD) report lower wanting for foods with high content of fat and protein unless they contain carbohydrates as well. A: Wanting ratings split by group and the macronutrients carbohydrates and fat. For display, we binned content so that low corresponds to the lower quartile, moderate to the second and third quartile, and high to the upper quartile. B: Differences between healthy control participants (HCP) and patients with MDD for foods according to their carbohydrate and fat content. Patients with MDD report lower wanting for high-fat foods (interaction contrast: *p* = .019). C: Wanting ratings split by group and the macronutrients carbohydrates and protein. D: Similar to fat, patients with MDD report lower wanting for high-protein foods (interaction contrast: *p* = .028).

The addition of protein instead of fat to the carbohydrate model led to a smaller improvement in model fit compared to modeling the carbohydrate and fat content for food wanting, *χ*^2^(11) = 292.73, *p* < .001, ΔBIC = 196 (with the carbohydrate model as reference). Increasing the protein content by 1 SD unit (∼6.0 g/100g) led to a reduction in wanting of *b* = -1.39 (*p* = .022) and this reduction was more pronounced in patients with MDD (*b* = -2.48, *p* = .042). A contrast comparing the slopes for carbohydrates and protein revealed a moderate relative preference for carbohydrates compared to fat in patients with MDD versus HCPs, *t*(117) = 2.22, *p* = .028. Analogous to liking, protein enhanced the wanting of carbohydrate-rich food, leading to a significant interaction term (*b* = 2.03, *p* < .001), and patients with MDD showed no differences compared to HCPs (*b* = 0.33, *p* = .68; Figure 2b; for item-level results, see Figure 3).

**Figure 3:**
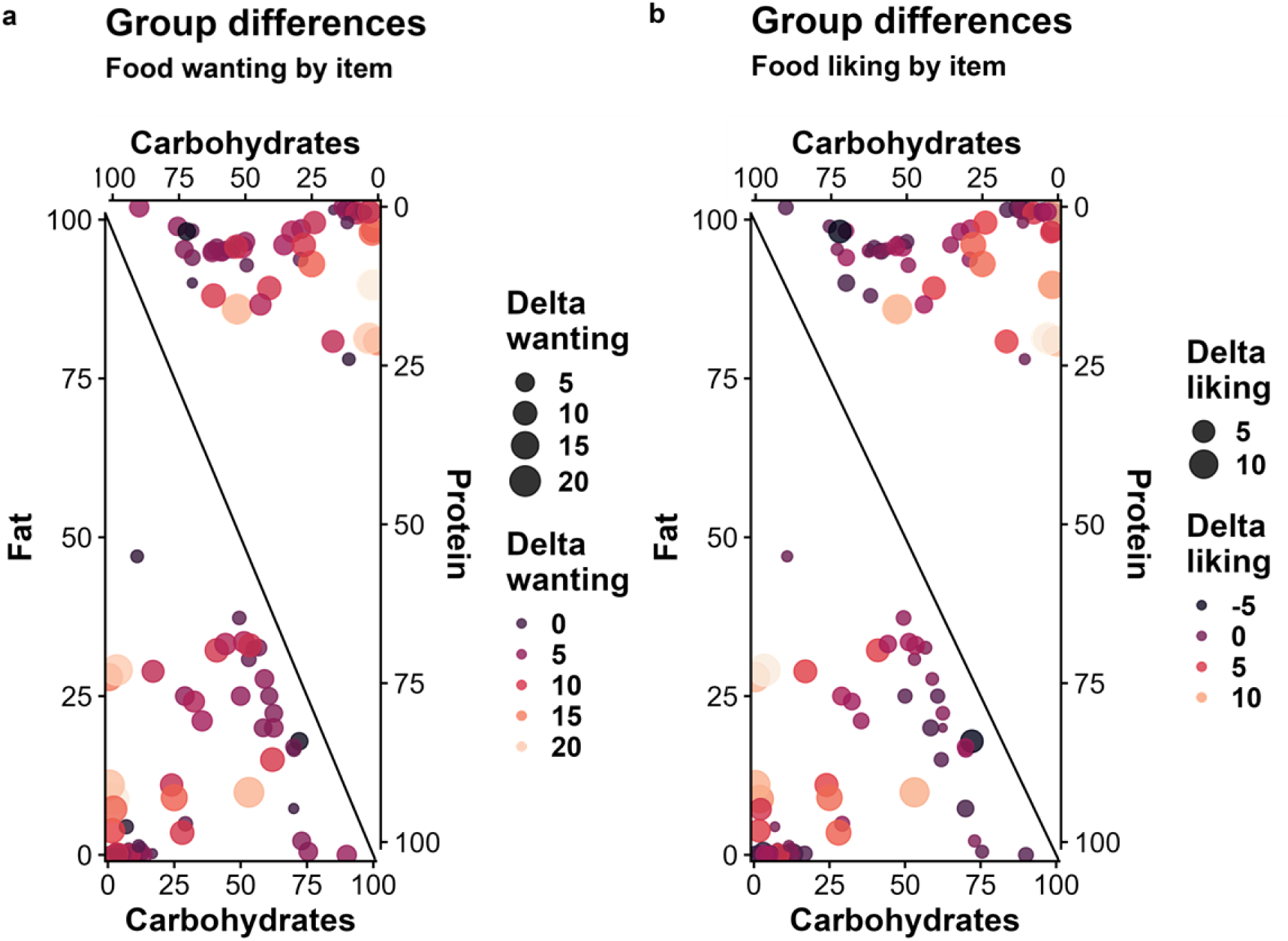
Differences in ratings between patients with major depressive disorder (MDD) and healthy control participants at the item level. A: Group differences in wanting ratings. The size of the dot corresponds to the absolute difference in ratings, whereas the color of the dot indicates the (signed) difference in ratings (higher values indicate larger deficits in patients). B: Group differences in liking ratings. For additional information on the selected items, see Table S1

### Patients with MDD show a reduced correlation between macronutrient slopes

To examine whether group differences in food ratings are driven by a disturbed correspondence between macronutrients, we ran additional analyses using unbiased individual estimates of effects (i.e., random slopes reflecting the increase of ratings by one unit of the macronutrient content) derived from modified linear mixed-effects models (see Methods). Notably, patients with MDD showed an attenuated association of individual carbohydrate estimates with fat estimates, *t*(113) = -3.78, *p* < .001, and an attenuated association with protein estimates (each predicted as outcomes by carbohydrate slope estimates), *t*(113) = -3.70, *p* < .001 (Figure 4). This difference in the correspondence between the macronutrient slopes suggests that patients with MDD have specific impairments in the metabolic sensing of fat and protein content while preferences for carbohydrate-rich foods are largely unaffected or even enhanced.

**Figure 4.**
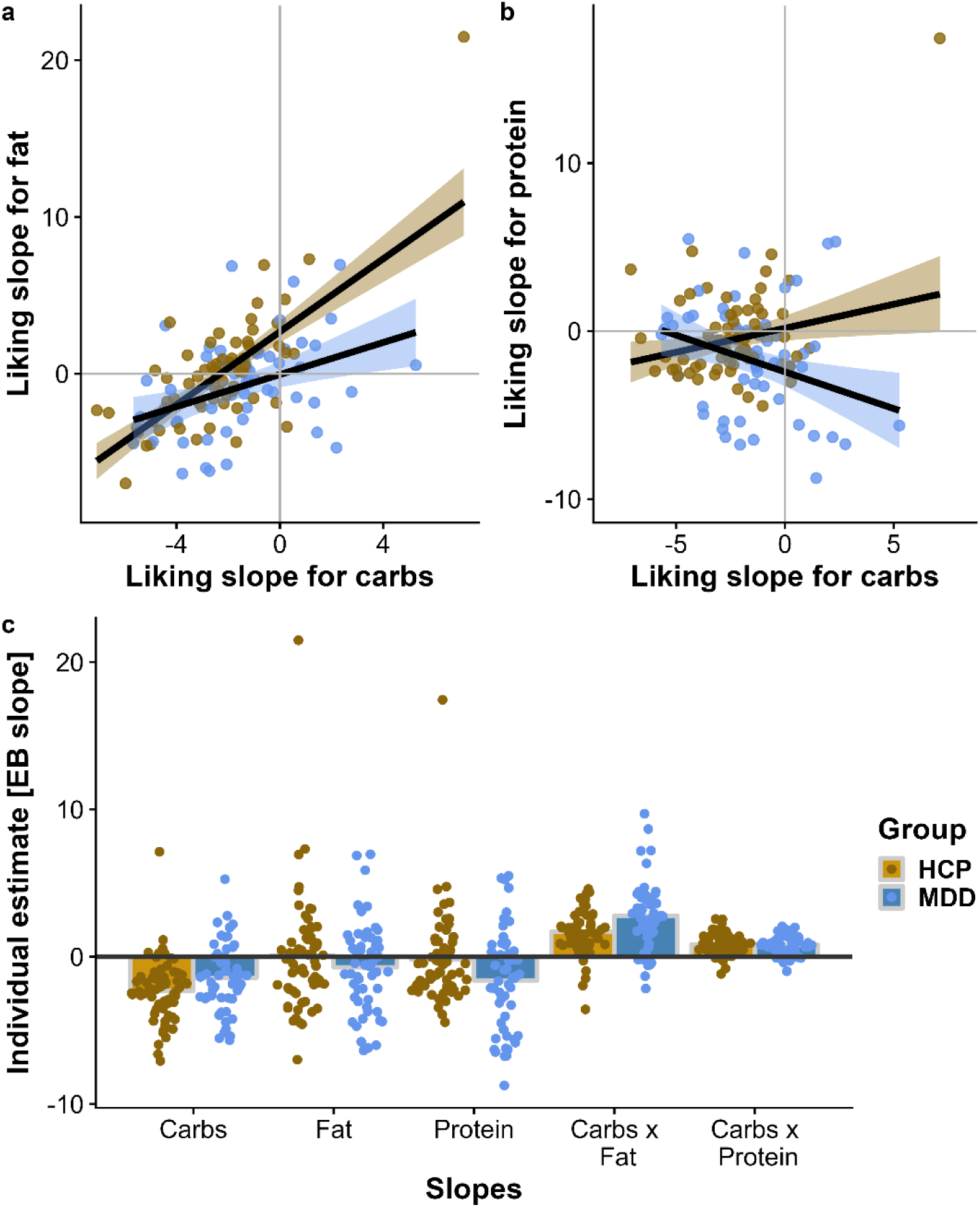
Patients with major depressive disorder (MDD) show attenuated associations between liking slopes for carbohydrates and fat as well as protein. A: Patients with MDD show a weaker correlation between individual estimates of increases in liking per one unit of carbohydrate content (i.e., liking slopes) and increases in liking for fat, *t*(113) = -3.78, *p* < .001. B: Patients with MDD show a negative correlation between liking slopes for carbohydrates and protein *t*(113) = -3.70, *p* < .001. C: Individual estimates per macronutrient split by group. The unbiased empirical Bayes (EB) estimates were derived from linear mixed-effects models that did not include group as a regressor. Patients with MDD show higher estimates in likings slopes for carbohydrates, protein, and Carbohydrates × Fat.

### Sensitivity and exploratory analyses

After establishing the associations of liking and wanting ratings with the macronutrient content of food, we conducted additional sensitivity analyses to explore associations of food liking ratings with severity, demographic, and metabolic variables (Figure 5). Participants with higher self-reported severity of depressive symptoms, as indexed by the BDI-II, reported lower overall liking (*r* = .197, *p* = .034) as well as higher liking for food rich in carbohydrates (*r* = .250, *p* = .007) and Carbohydrates × Fat (*r* = .302, *p* < .001). Similarly, participants with more severe anhedonia, as indexed by the SHAPS, reported lower overall liking (*r* = .277, *p* = .003) and higher liking for food rich in Carbohydrates × Fat (*r* = .204, *p* = .027). In contrast, changes in appetite and body weight, as indexed by the delta appetite score, did not correlate with any of the liking estimates. Additional follow-up analyses using the STAI-T to assess trait-like anxiety symptoms suggested robust associations between greater anxiety and liking of food rich in carbohydrates (*r* = .321, *p* < .001) and Carbohydrates × Fat (*r* = .352, *p* < .001).

**Figure 5.**
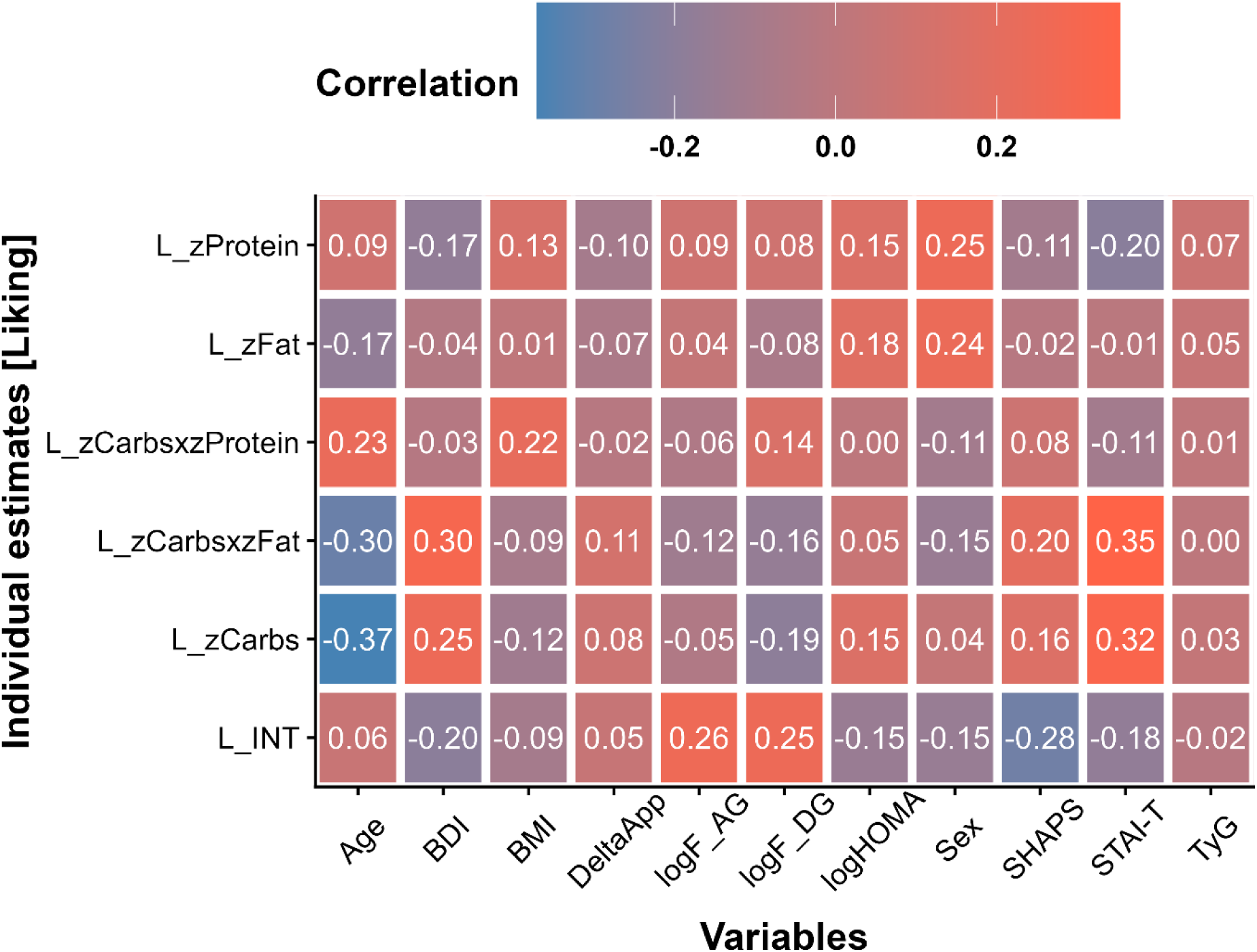
Results of the sensitivity analyses show associations with overall liking (L) and liking slopes reflecting associations of individual ratings with the macronutrient composition of the rated food items. INT = intercept, BDI = Beck Depression Inventory, BMI = body mass index, F = fasting, AG = acyl ghrelin, DG = desacyl ghrelin, HOMA = Homeostasis Model Assessment, SHAPS = Snaith-Hamilton Pleasure Scale, STAI-T = State-Trait Anxiety Inventory - Trait, TyG = Triglyceride-Glucose Index.

Whereas age, sex, and BMI were not significantly associated with overall liking, they showed correlations with macronutrient liking estimates. A higher BMI was associated with higher liking for food rich in Carbohydrates × Protein (*r* = .218, *p* = .018). A higher age was associated with lower liking ratings for food rich in carbohydrates (*r* = -.370, *p* < .001) and Carbohydrates × Fat (*r* = -.297, *p* = .001), but higher liking ratings for Carbohydrates × Protein (*r* = .234, *p* = .011). Men reported higher liking ratings for food rich in fat (*r* = .242, *p* = .009) or protein (*r* = .252, *p* = .006) compared to women.

Metabolic indices (transformed for parametric analyses) primarily showed associations with overall liking ratings. Both higher fasting levels of acyl (*r* = .257, *p* = .011) and desacyl ghrelin (*r* = .251, *p* = .013) were associated with higher overall liking ratings, whereas insulin sensitivity, as indexed by HOMA-IR and the triglyceride-glucose index, showed no significant associations with liking estimates. Associations with wanting slopes were qualitatively similar and the use of antidepressant medication did not affect the reported associations.

## Discussion

Reward dysfunction (anhedonia) is a cardinal symptom of MDD, and somatic symptoms (e.g., altered appetite) are common in severe depression. Although carbohydrate craving has long been recognized as a symptom of depression (Wurtman and Wurtman, 1995), a fully quantitative and experimental investigation of differences in perceived reward using the macronutrient composition of an extensive set of food items was missing to date. Using model comparisons, we demonstrate that the macronutrient composition of food explains substantial variance in individual ratings, providing an opportunity to dissect associations with individual estimates for mechanistic insights, bypassing recall-related biases of self-reported food intake (Ravelli and Schoeller, 2020). Consequently, we show that patients with MDD prefer carbohydrate-rich foods and experience food reward deficits for foods that are only rich in fat or protein while concurrently providing large amounts of carbohydrates compensate for these deficits. Crucially, the reward-enhancing effect of carbohydrate-rich foods was associated with the severity of depressive symptoms and anhedonia. Perhaps surprisingly, we observed no robust associations with the direction of changes in appetite that differentiate subtypes of depression. Instead, we found strong associations with trait-like symptoms of anxiety. These results suggest that preferences for carbohydrate-rich foods may play a bigger role in the regulation of anxiety- or mood-related symptoms (Christensen and Pettijohn, 2001), compared to somatic symptoms (i.e., delta appetite scores). This interpretation is supported by weak and non-significant associations of metabolic indices with individual estimates reflecting the effect of macronutrients on food reward ratings. To conclude, our results highlight the potential of considering the macronutrient composition of food to better understand candidate drivers of reward-related symptoms in MDD.

In accordance with clinical observations and earlier studies on depression (Rose et al., 2010; Paans et al., 2018; Paans et al., 2019), we observed a shift in liking and wanting ratings of patients with MDD towards foods rich in carbohydrates, compared to fat- or protein-rich foods alone. The significant interaction terms of carbohydrates with fat and protein further suggests that high carbohydrate content can compensate for the reduced perceived reward of foods rich in fat or protein. Notably, the estimated individual preferences for carbohydrates derived from the linear mixed-effects models (random slopes) were associated with symptom severity as indexed by the BDI-II, SHAPS, and STAI-T. Although our cross-sectional study design cannot address causality, preclinical experimental work has shown bidirectional links between dietary intake of carbohydrates and depressive symptoms. For example, a diet rich in refined carbohydrates facilitated anxiety and depressive-like behaviors after stress in mice (Santos et al., 2018) while a high-fat diet desensitizes the output of hypothalamic AgRP neurons, leading to dysregulation of behavior indicative of anxiety and depression (Xia et al., 2021). Moreover, the administration of citalopram, one of the commonly prescribed SSRIs, reduced the preference for fat in male, but not female rats (De la Fuente-Reynoso et al., 2022). In humans, dietary interventions seeking to reduce carbohydrate consumption have failed to induce robust antidepressive effects so far and, if anything, seem to increase anxiety, although the quality of the evidence is still low (Varaee et al., 2023). In contrast, recent Mendelian randomization studies even suggest a protective effect of carbohydrate consumption on the risk for depression (Yao et al., 2022). Therefore, longitudinal assessments of food liking and wanting in combination with interventions could be a highly promising approach to improve our understanding of the association between food preferences and depression.

At the mechanistic level, our results are mixed and highlight the necessity for follow-up work. On the one hand, reduced food reward ratings of patients with MDD for fat- and protein-rich foods support the idea that vagal afferent feedback signals might be dysregulated in depression (Tellez et al., 2013; de Araujo, 2016; de Lartigue and Diepenbroek, 2016; de Araujo et al., 2020; Goldstein et al., 2021). This interpretation is supported by the altered correlation between carbohydrate slopes and fat as well as protein slopes in patients with MDD derived from individual ratings of participants. On the other hand, metabolic indices reflecting insulin sensitivity were not associated with the estimated macronutrient slopes, and fasting levels of ghrelin were only associated with overall liking and wanting ratings. Although insulin sensitivity was robustly correlated with self-reported symptoms of anhedonia in our sample (Schulz et al., 2024), it showed no significant associations with food reward ratings. One explanation might be that insulin sensitivity has a stronger effect on the contrast between food and non-food rewards (Kroemer et al., 2013b; Tiedemann et al., 2017), which we did not evaluate here due to the focus on macronutrients. To resolve such open questions, interventional studies using intranasal insulin (Schneider et al., 2022) or ghrelin administration (Malik et al., 2008) in patients with MDD would be necessary.

Despite its notable strengths, such as the use of a representative set of well-known European foods that were independently rated in terms of liking and wanting leading to more than 7000 ratings per outcome, the study has several limitations that will need to be addressed in future work. First, due to the cross-sectional study design, it is not possible to answer whether food preferences for carbohydrate-rich foods in depression are functional (i.e., reducing symptoms) or dysfunctional (aggravating symptoms). An interventional design with longitudinal follow-up may help resolve this clinically highly relevant question. Second, we did not collect additional ratings to capture the assumed macronutrient composition of the depicted food items. It is conceivable that subjective evaluations beyond liking and wanting, such as estimated contents of fat or carbohydrates, would shed light on conscious versus subconscious processes related to food preferences (Tang et al., 2014). Relatedly, we collected no data on the typical frequency of consumption of the sampled foods, which might help elucidate potential links with eating behavior.

Depression is characterized by impaired reward sensitivity and opposing changes in appetite and body weight during depressive episodes. In line with previously reported carbohydrate cravings as a symptom of depression, we observed that patients with MDD show a marked preference for foods rich in carbohydrates, compared to foods rich in fat or protein. Moreover, the presence of carbohydrates in foods even compensated for the lower perceived reward in foods rich in fat in patients with MDD as indicated by the modeled interaction terms. Individual preferences for carbohydrates were further associated with depression severity, anhedonia, and trait-like anxiety symptoms, but unexpectedly not with the direction of changes in appetite during the current episode. Taken together, our study highlights that incorporating the macronutrient content of rated food items considerably improves our understanding of food reward deficits in depression and may pave the way for future interventional research. We anticipate that collecting liking and wanting ratings of food items could improve the longitudinal assessment of appetite-related symptoms compared to conventional food frequency questionnaires alone because it does not rely on potentially selective recall.

## Data Availability

All data produced in the present study are available upon reasonable request to the authors

## Acknowledgement

We thank Ebru Sarmisak, Nora Gerth, Yul Wegner, Anne Schiller, Antonia Schlaich, Johanna Voß, and Rauda Fahed for help with data acquisition. Stephanie Ebbinghaus kindly helped with running the ELISA tests. The study was supported by the German Research Foundation (DFG), KR 4555/7-1, KR 4555/9-1, KR 4555/10-1, and & WA 2673/15-1.

## CRediT Author contributions

**Lilly Thurn:** Formal Analysis, Visualization, Writing-Original draft preparation, Reviewing and Editing. **Corinna Schulz:** Visualization, Project administration, Investigation, Writing-Reviewing and Editing. **Diba Borgmann**: Writing-Reviewing and Editing. **Johannes Klaus**: Project administration, Investigation, Writing-Reviewing and Editing. **Sabine Ellinger**: Investigation, Writing-Reviewing and Editing. **Martin Walter**: Conceptualization, Methodology, Funding acquisition. **Nils B. Kroemer**: Conceptualization, Methodology, Funding acquisition, Supervision, Formal Analysis, Visualization, Writing-Original draft preparation, Reviewing and Editing.

## Financial disclosure

JK works as a study therapist in a multicenter phase IIb study by Beckley Psychtech Ltd on 5-MeO-DMT in patients with MDD, unrelated to this investigation. JK did not receive any financial compensation from the company. MW is a member of the following advisory boards and gave presentations to the following companies: Bayer AG, Germany; Boehringer Ingelheim, Germany; Novartis, Perception Neuroscience, HMNC and Biologische Heilmittel Heel GmbH, Germany. MW has further conducted studies with institutional research support from HEEL and Janssen Pharmaceutical Research for a clinical trial (IIT) on ketamine in patients with MDD, unrelated to this investigation. MW did not receive any financial compensation from the companies mentioned above. All other authors report no biomedical financial interests or other potential conflicts of interest.

## Notes

### Clinical Protocols

https://clinicaltrials.gov/study/NCT05318924

### Author Declarations

Ethics committee/IRB of University of Tuebingen, Faculty of Medicine gave ethical approval for this work

