## Supplemental Material for "Altered food liking in depression is driven by macronutrient composition"

Venusberg Campus 1, 53127 Bonn, Germany

**Table S1: Macronutrient composition of the selected foods**

| **Description** | **Energy_kcal_100g** | **TotalFat_g_100g** | **SaturatedFat_g_100g** | **Carbohydrate_g_100g** | **TotalSugar_g_100g** | **Protein_g_100g** | **Fiber_g_100g** |
| --- | --- | --- | --- | --- | --- | --- | --- |
| Milk chocolate | 546.0 | 32.5 | 20.7 | 54.9 | 49.0 | 6.9 | 2.8 |
| Biscuit with chocolate | 469.0 | 20.0 | 10.7 | 62.5 | 30.1 | 7.2 | 4.7 |
| Mars | 447.0 | 16.6 | 7.7 | 70.1 | 64.0 | 3.8 | 1.2 |
| Chocolate Cupcake iced with nuts | 548.4 | 20.0 | 10.1 | 58.5 | 38.0 | 7.0 | 0.9 |
| Bonbons (chocolates) | 519.0 | 30.8 | 16.8 | 53.1 | 51.8 | 5.6 | 3.3 |
| Whipped cream pie | 350.0 | 25.0 | 11.6 | 29.0 | 23.1 | 3.5 | 0.8 |
| Mini donuts | 358.0 | 21.1 | 4.2 | 35.5 | 4.8 | 6.0 | 1.1 |
| Kinder Bueno/Children's Bueno | 571.0 | 37.3 | 17.3 | 49.5 | 41.3 | 9.2 | 2.0 |
| Smarties 1 | 465.0 | 17.9 | 9.7 | 72.1 | 65.4 | 3.9 | 2.7 |
| Chocolate bar | 549.6 | 32.6 | 20.3 | 56.8 | 54.4 | 6.7 | 1.1 |
| Kitkat | 520.0 | 27.7 | 15.2 | 59.0 | 48.6 | 7.1 | 3.0 |
| Cake with chocolate | 450.0 | 25.0 | 14.8 | 50.0 | 32.4 | 5.5 | 2.3 |
| Chocolate chip cookies | 500.0 | 25.0 | 13.0 | 60.8 | 31.8 | 6.4 | 2.9 |
| Strawberry candies | 360.0 | 0.0 | 0.0 | 90.0 | 65.0 | 0.1 | 0.7 |
| Raisins | 326.0 | 0.5 | 0.2 | 75.5 | 72.8 | 3.1 | 3.7 |
| Strawberries with whipped cream | 75.6 | 4.4 | 2.9 | 6.9 | 6.7 | 0.9 | 0.9 |
| Pancakes | 196.0 | 4.9 | 2.1 | 29.2 | 4.9 | 8.3 | 0.6 |
| Strawberry pie with whipped cream | 205.0 | 11.0 | 4.7 | 24.0 | 13.0 | 2.5 | 1.7 |
| Apple (green) | 60.0 | 0.2 | 0.0 | 13.0 | 10.3 | 0.2 | 2.0 |
| Fruitsalad (berries, banana, melon) | 56.4 | 0.1 | 0.0 | 10.8 | 8.7 | 0.7 | 3.2 |
| Grapes, white | 78.0 | 0.2 | 0.1 | 16.8 | 16.2 | 0.5 | 1.7 |
| Tangerine | 45.0 | 0.1 | 0.0 | 9.8 | 8.2 | 0.7 | 0.9 |
| Kiwi, halves | 68.0 | 0.8 | 0.2 | 12.2 | 10.3 | 0.9 | 2.3 |
| Fruit salad | 45.7 | 0.2 | 0.0 | 9.5 | 8.7 | 0.4 | 1.3 |
| Nectarine | 33.0 | 0.1 | 0.0 | 6.5 | 6.5 | 1.0 | 1.1 |
| Orange | 51.0 | 1.0 | 0.2 | 7.8 | 7.7 | 0.8 | 2.0 |
| Cherries | 57.0 | 0.0 | 0.0 | 13.0 | 13.0 | 0.0 | 1.2 |
| Watermelon | 38.0 | 0.0 | 0.0 | 8.0 | 8.0 | 1.0 | 0.6 |
| Red berries | 48.0 | 0.0 | 0.0 | 5.0 | 4.0 | 1.0 | 8.2 |
| Black berries | 37.0 | 0.0 | 0.0 | 5.1 | 5.1 | 0.9 | 3.1 |
| Potato crisps (natural) | 541.0 | 33.5 | 5.9 | 51.3 | 0.5 | 6.4 | 4.1 |
| Cheese twist | 508.0 | 32.2 | 17.2 | 41.0 | 2.7 | 12.8 | 1.5 |
| Nuts | 577.0 | 47.0 | 6.9 | 11.0 | 3.5 | 24.0 | 7.0 |
| Crackers with medium mature white cheddar cheese | 415.0 | 28.9 | 15.6 | 17.0 | 2.0 | 21.2 | 0.6 |
| Nacho-cheese tortilla chips | 487.0 | 22.3 | 9.4 | 62.5 | 1.3 | 6.9 | 4.3 |
| Pepper potato crisps | 544.0 | 33.0 | 5.8 | 53.5 | 2.5 | 6.3 | 4.0 |
| Cheese plate | 338.0 | 28.0 | 20.3 | 0.3 | 0.3 | 21.2 | 0.0 |
| French fries with sauce | 367.7 | 24.1 | 15.5 | 32.4 | 1.0 | 3.9 | 2.7 |
| Cream cheese on toast | 370.3 | 9.9 | 8.1 | 53.1 | 2.5 | 16.1 | 2.7 |
| Spanish sausage | 358.0 | 29.1 | 11.3 | 3.4 | 3.0 | 20.7 | 0.0 |
| Cream crackers | 465.0 | 17.0 | 7.0 | 70.0 | 5.0 | 8.0 | 2.2 |
| Pretzels | 395.0 | 7.3 | 1.2 | 70.0 | 3.3 | 12.0 | 4.2 |
| Cocktail nuts | 547.0 | 33.2 | 7.6 | 44.3 | 4.6 | 15.4 | 4.8 |
| Bake rolls | 447.0 | 15.0 | 7.0 | 62.0 | 5.0 | 14.0 | 4.0 |
| Mini Snack a Jacks | 414.0 | 2.2 | 0.8 | 73.0 | 2.7 | 6.7 | 1.1 |
| Pizza bolognese slice | 220.0 | 9.0 | 3.1 | 25.0 | 2.0 | 9.0 | 2.0 |
| Pepper, yellow | 23.0 | 0.2 | 0.0 | 3.5 | 3.4 | 0.7 | 2.5 |
| Radish | 22.0 | 0.0 | 0.0 | 4.0 | 3.0 | 1.0 | 0.9 |
| Eggs, halves | 136.0 | 8.8 | 2.9 | 1.9 | 0.0 | 12.3 | 0.0 |
| Tomato, cut | 23.0 | 0.5 | 0.1 | 3.1 | 3.0 | 0.7 | 1.4 |
| Sweet pepper, red | 28.0 | 0.1 | 0.0 | 5.0 | 4.4 | 0.8 | 1.8 |
| Carrot | 33.0 | 0.3 | 0.1 | 5.5 | 4.3 | 0.6 | 2.8 |
| Cucumber | 13.0 | 0.2 | 0.1 | 1.9 | 1.7 | 0.6 | 0.6 |
| Sweet corn | 74.0 | 1.4 | 0.4 | 11.6 | 8.1 | 2.5 | 2.5 |
| Sushi | 165.0 | 3.5 | 0.1 | 28.0 | 6.3 | 6.0 | 0.8 |
| Greek salad | 92.0 | 7.2 | 3.1 | 2.2 | 0.6 | 4.0 | 1.1 |
| Mini peppers | 22.0 | 0.1 | 0.0 | 3.4 | 3.1 | 0.8 | 2.3 |
| Olive, pepper and cucumber salad | 57.9 | 3.8 | 2.8 | 1.7 | 2.0 | 3.6 | 1.2 |
| Mini pickles (sour) | 10.0 | 0.0 | 0.0 | 1.0 | 1.0 | 1.0 | 0.9 |
| Olives | 111.0 | 11.0 | 1.6 | 0.5 | 0.0 | 0.9 | 4.0 |
